# A protocol for a systematic review with prospective individual patient data meta-analysis in EGFR mutant NSCLC with brain metastases to assess the effect of SRS + Osimertinib compared to Osimertinib alone: the STARLET collaboration

**DOI:** 10.1101/2023.07.30.23293383

**Authors:** Kristy P Robledo, Shilo Lefresne, Yu Yang, Arjun Sahgal, Mark B Pinkham, Alan Nichol, Ross Andrew Soo, Ambika Parmar, Fiona Hegi-Johnson, Mark Doherty, Benjamin J Solomon, David Shultz, Ivan WK Tham, Adrian Sacher, Jeremy Tey, Cheng Nang Leong, Wee Yao Koh, Yiqing Huang, Yvonne Ang, Jiali Low, Clement Yong, Mei Chin Lim, Ai Peng Tan, Chee Khoon Lee, Cheryl Ho, the oSimertinib with or without sTereotActic Radiosurgery in egfr non-small cell Lung cancEr with brain metastases (STARLET) Collaboration

**Affiliations:** NHMRC Clinical Trials Centre, University of Sydney, Australia; BC Cancer Agency, Canada; National University Cancer Institute Singapore; Sunnybrook Health Science Centre, Canada; University of Queensland and Princess Alexandra Hospital, Australia; Sunnybrook Health Sciences Centre, Canada; Peter MacCallum Cancer Centre, Australia; St Vincents University Hospital, Ireland; Princess Margaret Cancer Centre, Canada; Mt Elizabeth Novena Hospital, Singapore; National University Hospital Singapore; National University Hospital, Singapore

**Keywords:** Individual participant data meta-analysis, EGFR mutant NSCLC, stereotactic radiosurgery, Osimertinib, brain metastases

## Abstract

**Background:** Patients with advanced non-small-cell lung cancer (NSCLC) with activating mutations in the epidermal growth factor receptor (*EGFR*) gene are a heterogenous population who often develop brain metastases (BM). The optimal management of patients with asymptomatic brain metastases is unclear given the activity of newer generation targeted therapies in the central nervous system. We present a protocol for an individual patient data prospective meta-analysis (IPD-PMA) to evaluate whether the addition of stereotactic radiosurgery (SRS) before Osimertinib treatment will lead to better control of intracranial metastatic disease. This is a clinically relevant question that will inform practice.

**Methods:** Randomised controlled trials (RCTs) will be eligible if they included: participants with BM arising from *EGFR* mutant NSCLC and suitable to receive Osimertinib both in the first- and second-line settings (P); comparisons of SRS followed by Osimertinib versus Osimertinib alone (I, C); and intracranial disease control included as an endpoint (O). Systematic searches of Medline (Ovid), Embase (Ovid), Cochrane Central Register of Controlled Trials (CENTRAL), CINAHL (EBSCO), PsychInfo, ClinicalTrials.gov and the World Health Organisation’s International Clinical Trials Registry Platform’s Search Portal will be undertaken. An IPD meta-analysis will be performed using methodologies recommended by the Cochrane Collaboration. The primary outcome is intra-cranial progression free survival, as determined by RANO-BM criteria. Secondary outcomes include overall survival, time to whole brain radiotherapy, quality of life and adverse events of special interest. Effect differences will be explored among pre-specified subgroups.

**Ethics and dissemination:** Approved by each trials ethics committee. Results will be relevant to clinicians, researchers, policymakers and patients, and will be disseminated via publications, presentations and media releases.

**Prospero registration:** CRD42022330532

**Strengths and Limitations of this study:**

- The use of an individual patient data (IPD) meta-analysis will give increased statistical power for the relative comparison of SRS followed by Osimertinib versus Osimertinib alone on intracranial progression-free survival. Such a meta-analysis will also enable the exploration of subgroups.
- Frequency of outcome assessment and outcome measures may be collected and reported differently across included trials, which may lead to some imprecision. Harmonisation of clinical trial protocols through prospective meta-analysis will address some of these limitations.
- A limitation of this study is that the searches will only be conducted until late 2023 and any studies that are registered after this time will not be included.

## INTRODUCTION

Non-small-cell lung cancer (NSCLC) with activating mutations in the epidermal growth factor receptor (*EGFR*) gene is a distinct subtype that is characterised by a high tumour response rate when treated with small molecule *EGFR* tyrosine kinase inhibitors (TKIs). Approximately 20% to 40% of patients with advanced NSCLC will develop brain metastases (BM) at some point during their disease course, and it is possible that patients with *EGFR* mutant NSCLC at greater risk due to improved survival (1, 2).

Stereotactic radiosurgery (SRS) involves the precise delivery of high doses of ionising radiation over a single or limited number of fractions to an intracranial target (3). Based on populations with BM from predominantly NSCLC but not enriched for *EGFR*, incorporating SRS in the management of BM was associated with improvement in overall survival (OS) for those with a single lesion and prolongation of functional independence in those with up to 3 BM (4). However, the detrimental effects of whole brain radiation are now well known such that SRS alone has become the standard of care. Use of SRS alone for multiple BM has been adopted routinely (5), in particular, given the prospective Japanese observational study involving patients with up to 10 BM demonstrated that OS of patients with 5-10 brain metastases treated with SRS alone was non-inferior to those with 2-4 brain metastases (6). Hence for patients with a good performance status, the American Society for Radiation Oncology (ASTRO) strongly recommends SRS for those with 1 to 4 BM and also conditionally recommends this treatment for those with 5 to 10 BM (7).

Osimertinib is an oral 3rd generation irreversible mutant selective, wild type sparing *EGFR* TKI with a higher central nervous system penetration and intracranial activity than first-generation *EGFR* TKIs. It has been approved by the U.S. Food and Drug Administration (FDA) as a first line treatment for *EGFR* mutant NSCLC based on the FLAURA trial(8, 9), as well as second line treatment for those who have developed a T790M mutation after exposure to first generation *EGFR* TKI based on the AURA 3 trial (10). In subset analyses, patients with stable, asymptomatic BM had significantly prolonged intracranial disease progression-free survival (ic-PFS) with Osimertinib compared to Geftinib or Erlotinib in the FLAURA trial (11) and platinum-pemetrexed in the AURA3 trial (12). However, the true intracranial activity of Osimertinib remains unclear as a significant number of patients enrolled in these trials had prior cranial radiotherapy (24% in FLAURA and 41% in AURA3). Notably, the OCEAN trial, a single arm phase two study of with T790M positive *EGFR* mutant NSCLC and untreated BM, found the intracranial response rate for second line Osimertinib was 67% and median ic-PFS was 25 months (13).

Currently, the optimal sequencing of SRS and Osimertinib in patients with *EGFR* mutant NSCLC and untreated BM is unclear. The American Society of Clinical Oncology (ASCO)-Society for Neuro-Oncology (SNO)-ASTRO guideline states that local therapy may be delayed in selective patients with asymptomatic BM from *EGFR* mutant NSCLC, however the strength of the recommendation is weak as the quality of evidence supporting this recommendation is low (14). There is conflicting evidence from retrospective cohort studies. Magnuson and colleagues found that those who received upfront cranial irradiation had longer OS than those who received upfront first-generation *EGFR* TKI with deferred cranial irradiation (15). Similarly, Yu and colleagues observed that upfront cranial radiotherapy was associated with reduced cumulative incidence of ic-PFS in the entire cohort receiving Osimertinib and improvement in OS in a subset of patients with 1-3 BM (16). However, Thomas and colleagues did not find any improvement (17).

Two phase II randomised controlled trials (RCTs), OUTRUN (TROG 17.02) (18) and LUOSICNS (19) are independently recruiting participants with BM from *EGFR* mutant NSCLC to evaluate whether SRS followed by Osimertinib is more efficacious than Osimertinib alone in delaying progression of intracranial disease. OUTRUN completed recruitment in September 2022, and LUOSICNS completed recruitment in April 2023. Both have a sample size of 40 participants, and individually lack the statistical power to formally compare differences between treatment arms. They are hypothesis-generating to inform planning of a future definitive phase III RCT.

Therefore, we have developed a collaboration, STARLET (oSimertinib with or without sTereotActic Radiosurgery in egfr non-small cell Lung cancEr with brain metastases), to prospectively conduct an individual patient data (IPD) meta-analysis of these RCTs to compare the effects of SRS followed by Osimertinib versus Osimertinib alone followed by deferred local cranial therapies on intracranial disease control in patients with BM from *EGFR* mutant NSCLC. The purpose is to establish which treatment strategy will lead to better control of intracranial disease, and if there are subgroups of patients that might benefit more from the combination treatment strategies.

## METHODS AND ANALYSIS

A systematic review and IPD meta-analysis will be conducted according to the recommended methods (20, 21). Lead investigators of eligible RCTs will be invited to share their IPD and join this STARLET collaboration. Online supplementary Appendix 1 lists eligible RCTs identified up to July 2022. This protocol adheres to the Preferred Reporting Items for Systematic Reviews and Meta-Analysis (PRISMA) extension for protocols (PRISMA-P, checklist detailed in supplementary Appendix 2) (22) and has been registered on PROSPERO (CRD42022330532). If subsequent potentially eligible RCTs are published, a nested prospective meta-analysis may be used, in order to combine retrospective inclusion of these additional trials with the proposed results gained from these analyses. At this time, there are no consumers actively involved with the collaboration.

### Eligibility criteria

#### Types of studies

STARLET will include RCTs only. Randomisation may occur at the individual level or by cluster and quasi-randomised trials will be excluded. There are no language or date restrictions.

#### Trial participants

Participants will be eligible if they are receiving Osimertinib in the first- or second-line setting. For those receiving Osimertinib as first line systemic therapy, all newly diagnosed participants must have a documented sensitising *EGFR* mutation (including exon 19 del, L858R (exon 21), G719X (exon 18), L861G (exon 21), S768I (exon 20) and T790M (exon 20)) and intracranial metastasis, with or without extracranial disease. For those receiving Osimertinib as second line systemic therapy, participants will have developed intracranial metastases while on first-line 1st or 2nd generation *EGFR* TKI therapy, with no or stable extracranial disease regardless of T790M mutation.

Intracranial disease is defined as: (a) ≤ 10 lesions visible and measurable on protocol screening MRI, with at least one BM amenable to SRS; (b) no single BM exceeding 30mm in longest diameter; and (c) absence of neurologic symptoms except for headache, nausea or seizure which were medically controlled.

#### Interventions

One intervention is SRS followed by Osimertinib. The SRS dose-fractionation schedule depends on size and location of the lesion. The SRS is to be planned after randomisation, and Osimertinib commences after the completion of SRS. Osimertinib treatment is described below.

The other intervention is Osimertinib alone. Osimertinib will be administered orally as one 80 mg tablet once a day. A cycle of treatment is defined as 28 days of once daily Osimertinib treatment.

For those allocated to Osimertinib alone, treatment with Osimertinib will commence following randomisation. Participants may continue to receive treatment with Osimertinib as long as they are continuing to show clinical benefit, as judged by the treating clinician, and within the guidelines of the relevant trial protocol’s discontinuation criteria.

### Information sources and search strategy

We searched the following databases from their inception: Medline (Ovid), Embase (Ovid), Cochrane Central Register of Controlled Trials (CENTRAL), CINAHL (EBSCO), PsychInfo, ClinicalTrials.gov and the World Health Organisation’s International Clinical Trials Registry Platform’s Search Portal. The full search strategy is available in **Online Supplementary Appendix 2**. The initial search was completed up to July 2022, and will be updated regularly to search for new trials until late-2023. Collaborators and contacts were asked to notify us of any additional planned, or ongoing completed trials that may fulfil eligibility criteria. At this time, only the two aforementioned trials (OUTRUN, TROG 17.02: NCT03497767 and LUOSICNS: NCT03769103) have been identified, and both trial teams have agreed to share IPD for this collaboration.

#### Selection of studies for inclusion in the review

Two members of the STARLET Collaboration will independently screen all future retrieved records against eligibility criteria. Any discrepancies will be resolved by consensus or, if required, adjudication by a third reviewer. The Principal Investigator and/or corresponding author of any additional eligible studies will be invited to join the STARLET Collaboration. If there is no response to initial emails, we will contact other co-authors or contacts listed on registration records. If IPD are not available for an eligible trial, we will use aggregate data where possible.

### Data collection, management, and confidentiality

#### Data receipt / extraction

De-identified IPD will be shared via secure data transfer platforms or via institutional secure email using password-protected zip files. Data will be provided according to a pre-specified coding template where possible, otherwise, data will be accepted in any format and recoded as necessary. The data management team will receive and store the data in perpetuity in a secure, customised database at the NHMRC Clinical Trials Centre, University of Sydney, and data management will follow the University of Sydney’s Data Management Policies. Each trial team will also be asked to provide metadata (such as questionnaires, data collection forms, and data dictionaries) to aid understanding of the datasets. Trial-level data, such as intervention details (setting, timing and duration), intervention details, method of sequence generation, allocation concealment, geographical location, sample size, outcome measures and definitions will be cross-checked against published reports, trial protocols, registration records and data collection sheets, in order to ensure data integrity.

#### Data processing

IPD from each trial will be checked with respect to range, internal consistency, consistency with published reports and missing items. Integrity of the randomisation process will be examined by reviewing the chronological randomisation sequence and pattern of assignment, as well as the balance of participant characteristics across intervention and control groups. Any inconsistencies or missing data will be discussed with trialists/data managers and resolved by consensus. Each included trial will be analysed individually, and results shared with trialists for verification. Once finalised, data from each of the trials will be combined into a single database.

### Risk of bias assessment and certainty of evidence appraisal

Included studies will be assessed for risk of bias by two reviewers, independently, using the criteria described in the Cochrane handbook (23): random sequence generation; allocation concealment; blinding of participants and personnel; blinding of outcome assessment; incomplete outcome data; selective reporting; and other bias. The quality of evidence will be assessed using the Grading of Recommendations Assessment, Development and Evaluation (GRADE) approach (24). Any differences will be resolved by consensus or with a third reviewer.

### Primary outcome

The primary outcome will be ic-PFS at 12 months, as determined by RANO-BM criteria (25).

### Secondary outcomes

All outcomes and their definitions are detailed in Table 1. Secondary outcomes include OS, time to whole brain radiotherapy, quality of life and adverse events of special interest (AESI).

**Table 1:** Outcomes for individual patient data meta-analysis.

| Outcome | Definition |
| --- | --- |
| **Primary outcome** |  |
| Intracranial progression free survival (iPFS) at 12 months | Time from randomisation to intracranial disease progression, as defined according to RAN-NO |
| **Secondary outcomes** |  |
| PFS | Time from randomization to any disease progression |
| Overall survival | Time from randomization to death |
| Time to salvage whole brain radiotherapy | Time from randomization to salvage whole brain radiotherapy |
| Time to salvage SRS | Time from randomization to salvage stereotactic radiotherapy |
| time to local brain failure | Time from randomization to local brain failure |
| time to distant progression | Time from randomization to distant disease progression |
| QOL | QLQ-C30 |
|  | QLQ-BN 20 |
| Adverse events of special interest | Rates of the following AE (for example): radiation necrosis, neurocognitive impairment, edema cerebral, muscle weakness right side, muscle weakness left side, Fatigue, Gait disturbance, Headache, Seizures, Tremors, Lethargy, Dizziness, Syncope, Stroke and Intracranial hemorrhage |

### Covariates and subgroups

Individual and study-level subgroup analyses will be conducted for ic-PFS. Individual-level characteristics to be assessed include mutation type (*EGFR* exon19 deletion vs exon 21 L858R vs uncommon sensitising mutations, pending numbers), line of therapy (first vs second), number of BM (either: <4 vs ≥4 *or* 1 vs ≥2, pending total numbers), diameter of largest lesion (≤15mm vs >15mm), age at baseline (<70 vs ≥70 years), sex (male vs female), country of treatment (Singapore vs Australia vs Canada), ethnicity (Asian vs other), smoker (never vs ex or current smoker), extracranial disease presence at baseline (present vs absent), and ECOG performance status (0 vs ≥1). If data are insufficient for the pre-specified subgroup analyses, categories will be reassessed prior to any analyses, by consensus of the STARLET collaboration.

### Data analysis

A detailed statistical analysis plan will be prepared and agreed on by the STARLET Collaboration members prior to any analyses being undertaken. Analyses will include all randomised participants who meet the inclusion criteria, for which IPD are available. All analyses will be based on randomised treatment allocation (intention to treat principle).

For the primary outcome of ic-PFS at 12 months, cumulative incidence estimates taking into account the competing risk of extracranial progression (with their variances) from the trials will be pooled using inverse variance weighting (two stage approach). Other secondary outcomes will be examined using Cox regression or linear models, adjusted for study (one stage approach). Heterogeneity of treatment effects across trials will be estimated using I, and investigated by fitting a trial-by-treatment interaction term to the models. Any heterogeneity identified will be explored further.

Differences in treatment effect between the pre-specified subgroups will be examined by testing a treatment by subgroup interaction term within a Fine-Gray regression model for ic-PFS, taking into account competing risks. Findings of subgroup analyses will be reported as exploratory.

Missing data may be explored in sensitivity analyses using multiple imputation. Analyses will be performed using SAS and the open-source software R (26).

### Assessment of selection or publication bias

Potential selection bias and publication bias may be investigated by conducting a nested prospective meta-analysis and comparing trials that were included prospectively versus those identified retrospectively in a sensitivity analysis (if appropriate). Contour-enhanced funnel plots to examine whether there are differences in results between more and less precise studies.

### Adjustments for multiple testing

No formal adjustments will be made for multiple comparisons. However, we will follow Schulz and Grimes’ approach (27) and interpret the patterns and consistency of results across related outcomes rather than focusing on statistical significance alone.

### Planned sensitivity analyses

If possible, the following sensitivity analyses will be conducted for the primary outcome; including published aggregate data combined with IPD in the meta-analysis compared to IPD alone and including prospectively included trials only. Additional sensitivity analyses may be conducted on other outcomes to determine the effect of missing data. These will be detailed in the statistical analysis plan.

### Project management

Membership of the STARLET Collaboration includes representatives from each of the trials contributing IPD to the project. Trial representatives have the opportunity to contribute their expert knowledge to the Collaboration and provide input into the protocols, statistical analysis plan, and final results manuscript. The STARLET Collaboration will be responsible for data collection, management and analysis, as well as communication within the Collaboration, including organisation of virtual or face-to-face collaborator meetings.

## ETHICS AND DISSEMINATION

### Ethical considerations

IPD will be provided by each included trial on the stipulation that ethical approval has been provided by their respective Human Research Ethics Committees (or equivalent), and participants gave informed consent before enrolment. Only trials with ethics approval will be included in these analyses. Trialists remain the custodians of their own data, which will be de-identified before being shared with the Collaboration.

### Publication policy

Manuscripts will be prepared by the relevant members of the STARLET Collaboration, and circulated for comment, revision and approval prior to submission for publication. Any reports of the results from this study will be published either in the name of the collaborative group, or by representatives of the collaborative group on behalf of the STARLET Collaboration, as agreed by all members.

## DISCUSSION

An IPD meta-analysis is considered the gold standard of systematic reviews, and has many advantages over a standard aggregate approach. This includes the collaboration of a range of expert trialists and biostatisticians in order to ensure that all possible RCTs are included and appropriate analysis of outcomes are performed. Through prospectively collaborating, STARLET can pre-specify the patient population, interventions, and outcomes clearly and harmonise trial protocols where possible. Another clear advantage is the increase in statistical power. The two eligible RCTs identified at this time are both Phase II RCTs that individually are not powered to identify a statistically significant difference between treatments, but rather are looking for efficacy signals and safety of treatment.

We will seek to address this with the use of a prospective meta-analysis to include published aggregate data, and by encouraging planned and ongoing trials to collect our core outcomes and share data.

Based on the recruitment timelines of the two trials identified, we plan to complete study identification by end of 2023, IPD collection by mid-2024 and conduct the analyses and disseminate the results by mid-2025. These timelines may be adjusted if follow-up completes early, or if additional trials are identified and not completed in time to provide data.

The results of this systematic review will guide whether a Phase III study is required to inform clinical practice, and, if so, may help investigators to pre-plan subgroup analyses of interest.

## Supporting information

Supplementary 1 (search)

Supplementary 1 PRISMA

## Data Availability

Data sharing is not applicable to this article as no datasets were generated or analysed during the current study.

## DECLARATIONS

### Ethics approval and consent to participate

Each trial contributing data to this project has been approved by ethics committees. OUTRUN (TROG 17.02, NCT03497767) has ethics approval from Melbourne Health Human Research Committee, Melbourne, Australia and LUOSICNS (NCT03769103) has ethics approval from British Columbia Cancer Agency Research Ethics Board, University of British Columbia, Canada.

### Consent for publication

NA

## Authors Contributions

KR, CKL, YY, FHJ, CH, AN and SL conceived the idea for this study. KR, YY, CKL, FHJ, MP, RS, CH, AN and SL developed the research question and protocol registration. KR wrote the first draft of the manuscript. KR and YY developed the eligibility and search strategy, and performed the search and screening. All authors critically reviewed and provided feedback on the intellectual content of the manuscript and agreed and approved the final version.

## Acknowledgements

We would like to thank AstraZeneca for their ongoing support of both clinical trials. We would also like to thank Cancer Australia for their support for OUTRUN.

YYS: Supported by the National Medical Research Council (NMRC/MOH/000396)

## Abbreviations

AESI: adverse events of special interest
ASCO: American Society of Clinical Oncology
ASTRO: American Society for Radiation Oncology
BM: brain metastases
*EGFR*: epidermal growth factor receptor
FDA: U.S. Food and Drug Administration
GRADE: Grading of Recommendations Assessment, Development and Evaluation ()
ic-PFS: intracranial disease progression-free survival
IPD: individual patient data
NSCLC: Non-small-cell lung cancer
OS: overall survival
PRISMA-P: Preferred Reporting Items for Systematic Reviews and Meta-Analysis (PRISMA) extension for protocols (P)
RCT: randomised controlled trial
SNO: Society for Neuro-Oncology
SRS: Stereotactic radiosurgery
STARLET: oSimertinib with or without sTereotActic Radiosurgery in egfr non-small cell Lung cancEr with brain metastases
TKI: tyrosine kinase inhibitors

