## Supplementary 1 (search) for "A protocol for a systematic review with prospective individual patient data meta-analysis in EGFR mutant NSCLC with brain metastases to assess the effect of SRS + Osimertinib compared to Osimertinib alone: the STARLET collaboration"

STARLET: Supplementary One

Search strategies

The following search strategy will be amended for the relevant database. The example given below is for Medline via OVID.

Database: Ovid MEDLINE(R) ALL <1946 to June 07, 2022>

Search Strategy:

--------------------------------------------------------------------------------

1	Carcinoma, Non-Small-Cell Lung/ (64897)

2	lung cancer.mp. (184784)

3	(lung metast* or lung tumo* or lung carcinoma*).mp. (58213)

4	1 or 2 or 3 (230163)

5	cerebral ventricle neoplasms/ or infratentorial neoplasms/ or supratentorial neoplasms/ (6722)

6	(brain metast* or brain carcinom* or cerebral metast* or intracranial metast*).mp. (17732)

7	5 or 6 (24339)

8	ErbB Receptors/ (45155)

9	(egfr* or epidermal growth factor receptor*).mp. (95498)

10	8 or 9 (103850)

11	4 and 7 and 10 (915)

12	Protein Kinase Inhibitors/ (54970)

13	Antineoplastic Agents/ (310465)

14	(Osimertinib or Tagrisso or AZD9291).mp. (2096)

15	12 or 13 or 14 (351853)

16	11 and 15 (401)

17	cranial irradiation/ or radiosurgery/ or radiotherapy, computer-assisted/ or radiotherapy, high-energy/ or radiotherapy, image-guided/ or x-ray therapy/ (40233)

18	radiosurg*.mp. (24054)

19	(radioherap* or radiation or irradiation).mp. (759833)

20	17 or 18 or 19 (777639)

21	16 and 20 (114)

22	randomized controlled trial.pt. (570102)

23	controlled clinical trial.pt. (94897)

24	randomized.ab. (564115)

25	clinical trials as topic.sh. (200057)

26	randomly.ab. (384145)

27	trial.ti. (264006)

28	22 or 23 or 24 or 25 or 26 or 27 (1403243)

29	exp animals/ not humans.sh. (5015432)

30	28 not 29 (1294353)

31	21 and 30 (17)

Review of these 17 trials gave two eligible for inclusion at the time of this search.

OUTRUN:	A Randomised Phase II Trial of Osimertinib With or Without SRS for EGFR Mutated NSCLC With Brain Metastases. <https://ClinicalTrials.gov/show/NCT03497767>.

LUOSICNS:	Study of Osimertinib + SRS vs Osimertinib Alone for Brain Metastases in EGFR Positive Patients With NSCLC. <https://ClinicalTrials.gov/show/NCT03769103>.
